# QUALITY OF LIFE AND ITS DETERMINANTS AMONG INFERTILE AND NON-INFERTILE WOMEN: A CASE-CONTROL STUDY IN GANDAKI PROVINCE, NEPAL

**DOI:** 10.1101/2024.01.23.24301664

**Authors:** Rajendra Regmi, Dipendra Kumar Yadav, Sirjana Tiwari

## Abstract

**Introduction:** Infertility is highly stressful to married couple and has various social and psychological problems leading adverse impact on quality of life. The study examined the quality of life and factors associated with quality of life among infertile and non-infertile women.

**Materials and Methods:** Case control study was carried out among infertile and non-infertile women to compare their QoL using the World Health Organisation Quality of Life-BREF (WHOQOL-BREF) questionnaire. Altogether 92 married women of reproductive age 20-49 facing infertility/subfertility problems were selected as cases and controls were selected in 1:1 ratio with cases after matching.The written and verbal inform consent was taken from patients and ethical approval was taken from NHRC. Epi-data was used for data entry and data was analyzed using SPSS. The data collection in this study was from May 20 2019 to June 20 2019. Multivariable analysis was applied to the variable after bivariate analysis for the adjustment.

**Results:** The prevalence of infertility was found 9.1%, among then 43.5% had primary and 56.5% secondary infertility. The mean age of marriage of infertile women was significantly higher than that of non-infertile women (p 0.001).The average BMI score of infertile women was significantly higher than that of non-infertile women (p 0.001). Similarly the average perceived stress score among infertile women (28.9±4.61) and non infertile women (25.27±3.36), average anxiety score among infertile women (8.71±3.0) and among non-infertile (7.78±2.89), and average depression score among infertile women (8.14±2.67) and among non-infertile (6.86±2.49) were significantly higher in infertile women than non-infertile women. The total and subscale wise perceived social support score of infertile women was significantly lower than non-infertile women (p<0.001). The overall and inter-domain QoL score of infertile women was significantly lower than non-infertile women (p<0.001). Family planning methods used before first child (AOR-16.59, p=0.025), occupation (AOR-16.88, p=0.023) and induced abortion (AOR-0.086, p=0.047) were found as significant determinants of infertility at 95% CI. Among infertile women, only two factors, perceived stress (AOR-10.13, 95% CI: 3.52-29.18) and perceived social support (AOR-3.412, 95% CI: 1.15-10.101) found as important determinants of quality of life among infertile women, where as moderate to severe level of depression (AOR-14.61, 95% CI: 2.37-89.96); mild level of depression (AOR-3.42, 95% CI: 1.08-10.86), perceived social support (AOR-4.94, 95% CI: 1.51-16.14) and RH problems (AOR-3.539, 95% CI: 1.01-12.46) found as the determinants of quality of life among control (non-infertile women).

**Conclusion:** The findings of this study revealed that the overall and inter domain quality of life of infertile women were lower than that of non-infertile women. A community-based and multicultural study involving more districts may shed more light on this topic in future research. Health service strengthening, priority to infertility in RH programs and counseling sessions should be incorporated as part of the holistic approach in the day-to-day management of the infertile women.

## Introduction

Infertility in this study was taken as women not able to become pregnant and not maintaining that pregnancy to live birth and in male, when motile and viable sperm can’t find in ejaculation of man than such a male can be define as infertile. If very few viable sperms exist and have no zero risk of pregnancy in his female partner, than the male can be defined as sub-fertile or infertile. Now a days, through different treatment procedures and advances in reproductive technologies, assisted pregnancy is possible in the world, so patient friendly term “subfertility” is used as equivalent term of infertility.^1^

## MATERIALS AND METHODS OF STUDY

A a case control study was conducted after doing **s**creening survey among 1351 married women of reproductive age (20-49 years) to identify cases (infertile women) and controls (non-infertile women) from different four local institutions of Syangja and Kaski district of Gandaki Province, Nepal, which were selected on the basis of multistage sampling. The study population of this study was reproductive age (20-49 years) women facing infertility problems as case and reproductive age (20-49 years) women from same geographic area without infertility condition were selected as control. Selected sample size was 92 cases and 92 controls. Survey was conducted to find out the participants through screening questionnaires. Women of reproductive age at risk of becoming pregnant, who report unsuccessfully trying for a pregnancy for more than two years, was selected as cases and married women of reproductive age (20-49) without infertility problems from same geographical areas were included as control in the research study.Those clients who did not want to take part in research study and Known male factor infertility was excluded from the study. The controls were selected by matching age and level of literacy with cases. Written and verbal consent was taken from participants before data collection. Ethical clearance was be taken from Nepal Health Research Council. Semi structure questionnaire was used to collect the data using the World Health Organisation Quality of= Life-BREF (WHOQOL-BREF) questionnaire.The pretesting of individual tools for its validity and reliability was done in Pokhara 30 Khudi among 9 case and 9 controls. Epi-data was used for data entry and data was analyzed using SPSS. The data collection in this study was from May 20 2019 to June 20 2019. Multivariable analysis was applied to the variable after bivariate analysis for the adjustment.

## CHAPTER-IV - RESULTS

### 4.2.1 Socio-demographic characteristics of the participants

**Table 1:**
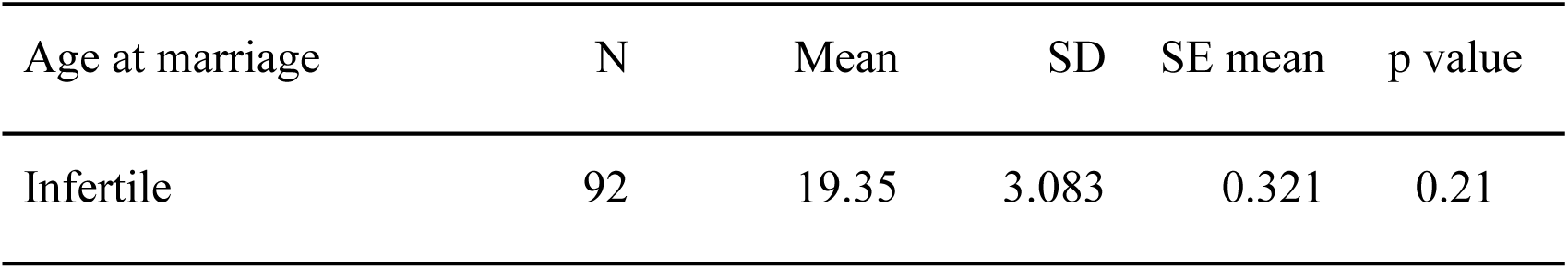
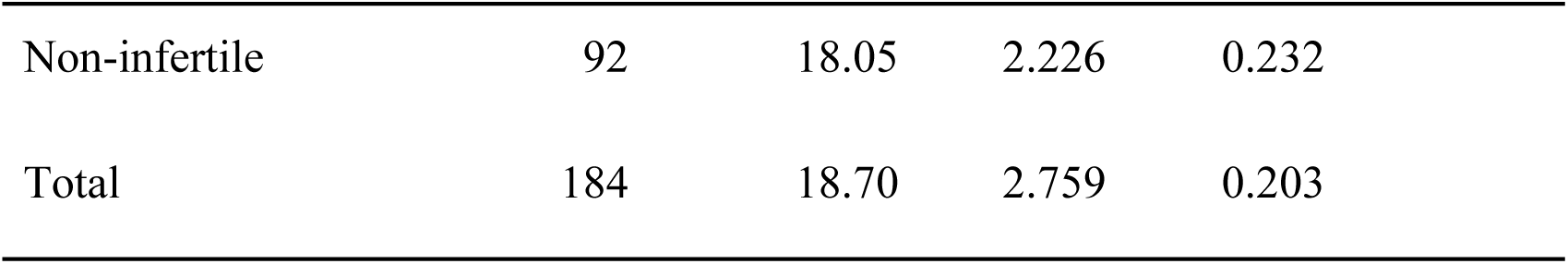
Age at marriage.

| Age at marriage | N | Mean | SD | SE mean | p value |
| --- | --- | --- | --- | --- | --- |
| Infertile | 92 | 19.35 | 3.083 | 0.321 | 0.21 |

|  |  |  |  |  |
| --- | --- | --- | --- | --- |
| Non-infertile | 92 | 18.05 | 2.226 | 0.232 |
| Total | 184 | 18.70 | 2.759 | 0.203 |

In this study 92 infertile/sub-fertile and 92 non-infertile women with the mean age at marriage was 32.86±6.201 were evaluated. The average (Mean±SD) age at marriage among infertile and non-infertile was 18.7±2.759 years. The mean age at marriage in infertile women was more than non-infertile ones, but the difference was not significant (p=0.21).

**Table 2:**
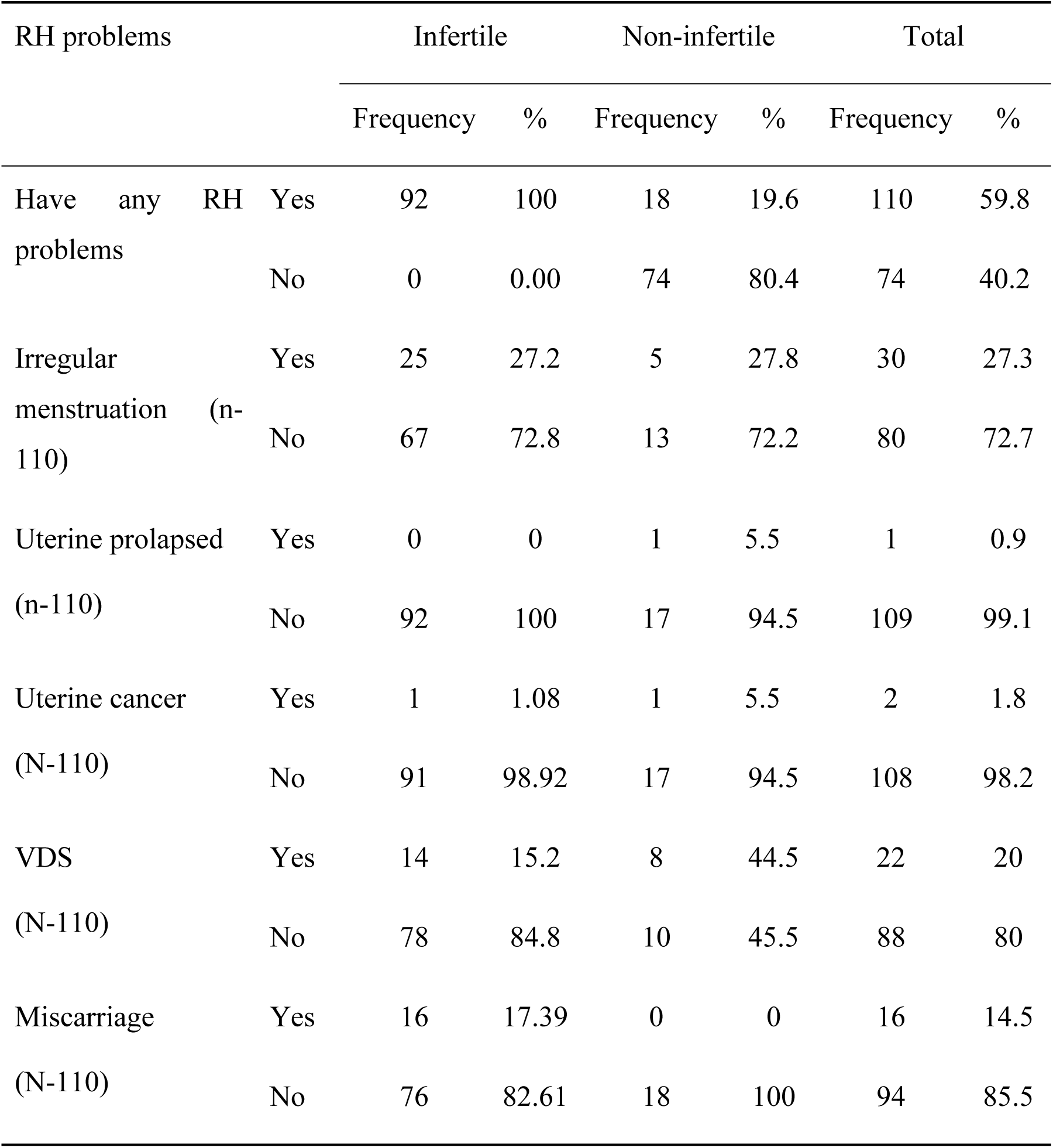
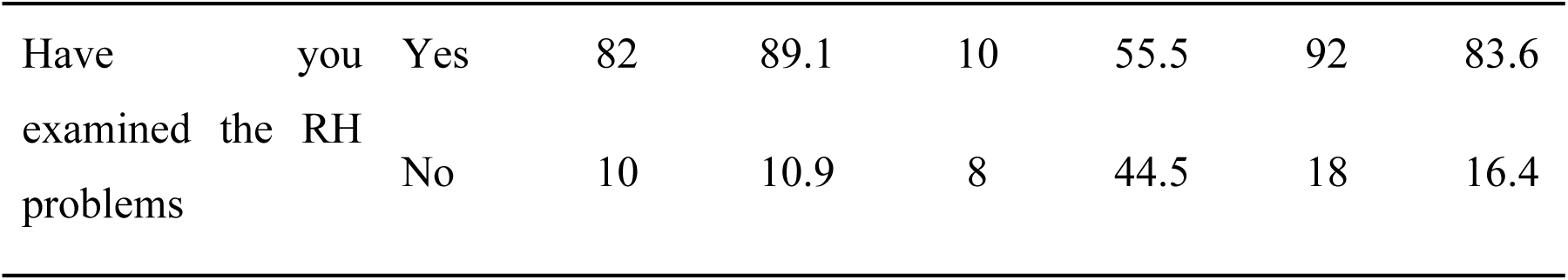
Reproductive problems.

Among total respondents 58.7% reported different reproductive health problems including infertility/sub-fertility during study, among them27.3% respondents were suffering from irregular menstruation, 20% from VDS, followed by miscarriage (14.5%), uterine cancer (1.8) and others.

19.6% of non-infertile women had reported different RH problems. The prevalence of irregular menstruation among infertile found 27.2%, whereas only 5.4 % among non-infertile. Similarly among women with reproductive health problems, 15.2% infertile women and 44.5% non infertile women were suffering from VDS. Among infertile women 81.5% had visited for examination treatment of RH problems, whereas 55.5 non-infertile women with RH problems found visited for examination/ treatment of RH problems

### 4.2.3 Psychosocial problems of the participants

Among different psychosocial problems the perceived stress, anxiety, depression, and social support of participants were measured using different tools in this study.

The study showed that the infertile women were perceived high stress, anxiety and depression than non-infertile women. Similarly infertile women found perceived low social support than non-infertile women.

**Table 3:** Psychosocial problems.

| Psychosocial problems |  | Infertile (n=92) |  | Non-infertile (n=92) |  | Total (n=184) |  |
| --- | --- | --- | --- | --- | --- | --- | --- |
|  |  | Frequency | % | Frequency | % | Frequency | % |
| Perceived stress |  |  |  |  |  |  |  |
| Below mean(normal stress) |  | 26 | 28.3 | 57 | 62.0 | 83 | 45.11 |
| Above mean(high stress) |  | 66 | 71.7 | 35 | 38.0 | 101 | 54.89 |
| Mean±SD |  | 28.93±4.62 |  | 25.27±3.57 |  | 27.103±4.50 |  |
| Anxiety and depression |  |  |  |  |  |  |  |
| Depression | Normal | 36 | 39.1 | 60 | 65.2 | 96 | 52.12 |
|  | Mild | 40 | 43.5 | 23 | 25 | 63 | 34.24 |
|  | Moderate | 15 | 16.3 | 9 | 9.8 | 24 | 13.04 |
|  | Severe | 1 | 1.1 | 0 | 0 | 1 | 0.5 |
|  | Mean±SD | 8.14±2.67 |  | 6.86±2.49 |  | 7.50±2.65 |  |
| Anxiety | Normal | 31 | 33.7 | 47 | 51.1 | 78 | 42.39 |
|  | Mild | 38 | 41.3 | 30 | 32.6 | 68 | 36.95 |
|  | Moderate | 20 | 21.7 | 13 | 14.1 | 33 | 17.93 |
|  | Severe | 3 | 3.3 | 2 | 2.2 | 5 | 2.72 |
|  | Mean±SD | 8.71±3.05 |  | 7.78±2.89 |  | 8.25±3.00 |  |
| Perceived Social Support |  |  |  |  |  |  |  |
| Other support | Low PSS | 1 | 1.1 | 0 | 0 | 1 | 0.5 |
|  | Medium PSS | 35 | 38 | 15 | 16.3 | 50 | 27.17 |
|  | High PSS | 56 | 60.9 | 77 | 83.7 | 133 | 72.28 |
| Family support | Low PSS | 5 | 5.4 | 0 | 0 | 5 | 2.72 |
|  | Medium PSS | 37 | 40.2 | 19 | 20.7 | 56 | 30.43 |
|  | High PSS | 50 | 54.3 | 73 | 79.3 | 123 | 66.85 |
| Friend | Low PSS | 3 | 3.3 | 2 | 2.2 | 5 | 2.72 |
| support | Medium PSS | 68 | 73.9 | 41 | 44.6 | 109 | 59.24 |
|  | High PSS | 21 | 22.8 | 49 | 53.3 | 70 | 38.04 |
| Total | Low PSS | 1 | 1.1 | 1 | 1.1 | 2 | 1.08 |
|  | Medium PSS | 56 | 60.9 | 19 | 20.7 | 75 | 40.76 |
|  | High PSS | 35 | 38 | 72 | 78.3 | 117 | 63.59 |
|  | Mean±SD | 58.77±7.87 |  | 64.06±7.78 |  | 62.22±8.54 |  |

Among infertile women 71.7% perceived high stress, where only 38% of non-infertile women were perceived high stress. Mean score of perceived stress among infertile and non-infertile was 27.103±4.50. In an average the infertile women found to perceived more stress (28.9±4.619) than non-infertile women (25.27±3.567).

Level of Anxiety and depression of participants were measured using HADS tools. Among infertile women, 39.1% had normal depression level, 43.5% mild depression level among, 16.3% had moderate depression level and only 1.1% infertile women had severe depression level. Similarly among non-infertile women nearly two thirds (65.2%) of non-infertile women had normal depression level, 25% had mild level and nearly one in ten (9.8%) had moderate level of depression.

One third of infertile women (33.7%) had normal anxiety level, 41.3% had mild, 21.7% had moderate and only in 3.3% infertile women found severe anxiety level. Similarly among non-infertile women more than half (51.1%) respondents had normal anxiety level, nearly one third (32.6%) had mild anxiety level, moderate level anxiety problem in 14.1% and severe level of anxiety among 2.2% of non-infertile women.

Among infertile women 1.1% were perceived low social support nearly two third (60.09%) infertile women found perceived medium social support and more than one third (38.0%) of infertile women perceived high social support. Similarly among non-infertile women only 1.1% had perceived low social support, 20.7% had medium social support and 78.3% perceives high social support.

**Table 4:** Quality of life score of respondents.

| Domain | Infertile n=92 |  | Non-infertile<br>n=92 |  | Total n= 184 |  |
| --- | --- | --- | --- | --- | --- | --- |
|  | Mean | SD | Mean | SD | Mean | SD |
| Physical | 60.66 | 9.38 | 68.64 | 11.16 | 64.65 | 11.03 |
| Psychological | 50.85 | 9.92 | 57.64 | 9.61 | 54.25 | 10.32 |
| Social Relations | 57.97 | 12.03 | 67.75 | 13.37 | 62.86 | 13.60 |
| Environment | 48.64 | 10.37 | 57.98 | 10.46 | 53.31 | 11.39 |
| Overall score | 54.01 | 7.79 | 62.30 | 7.82 | 58.15 | 8.82 |

From above table the mean quality of life score among total respondents was 58.15±8.82. Average total quality of life score as well as domain wise quality of life score among infertile women (54.01±7.79) was lower than that of non-infertile women (62.3±7.82).

### 4.3 Bivariate Analysis

#### 4.3.1 Factors associated with infertilities

**Table 5:**
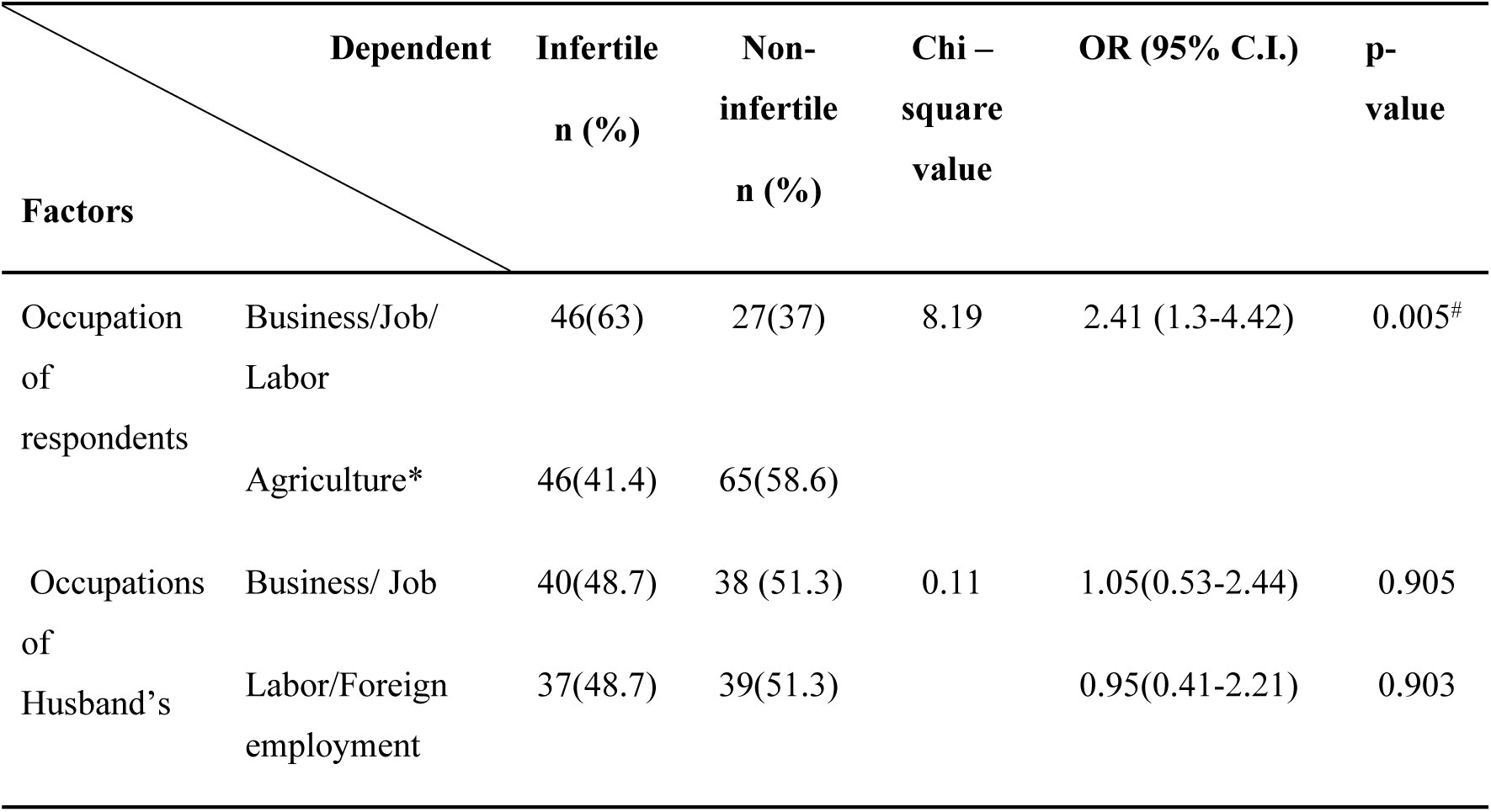
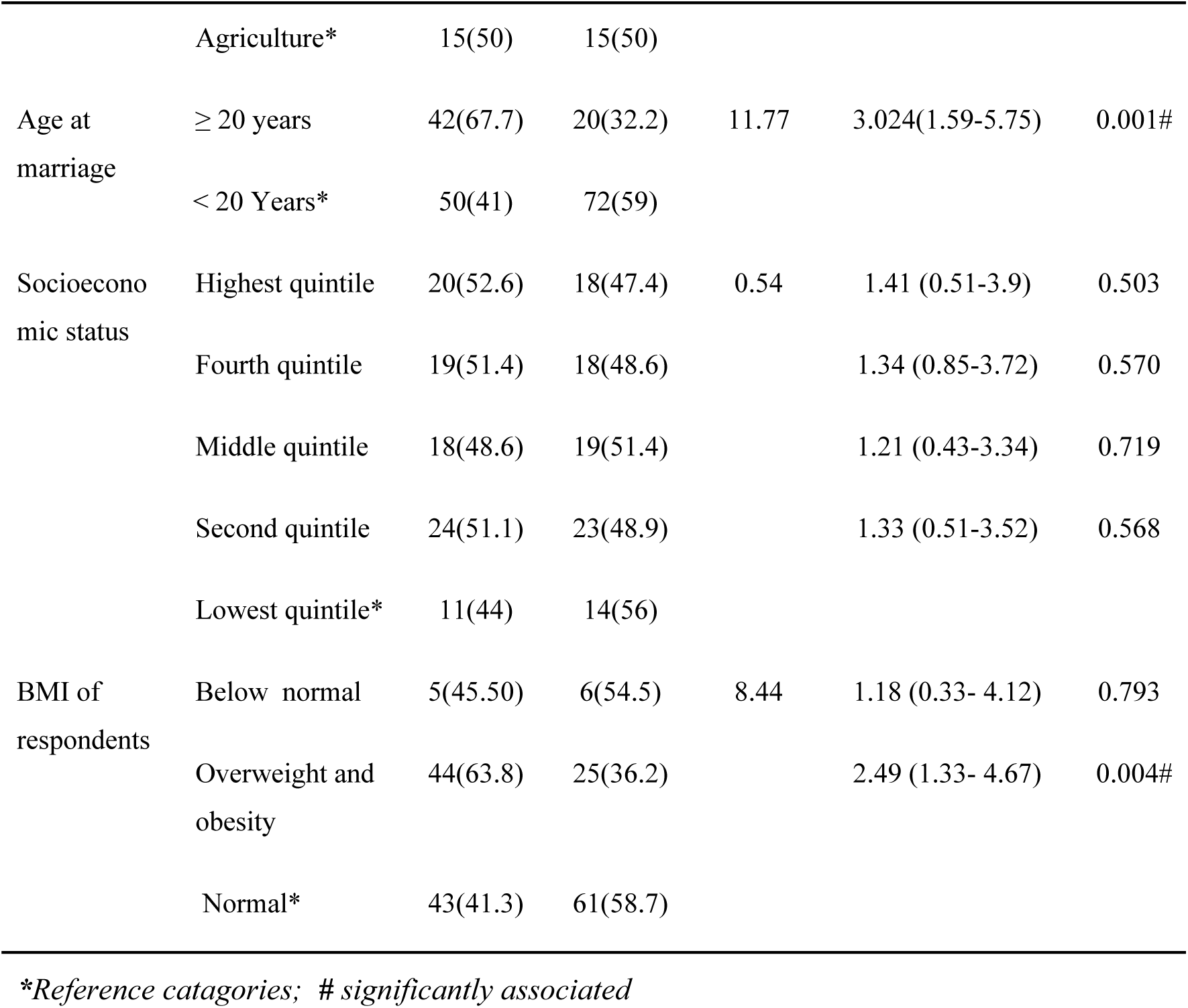
Association of infertility problems with demographic characteristics.

| Factors |  | Dependent | Infertile<br>n (%) | Non-infertile<br>n (%) | Chi –<br>square<br>value | OR (95% C.I.) | p-value |
| --- | --- | --- | --- | --- | --- | --- | --- |
| Occupation<br>of<br>respondents | Business/Job/<br>Labor |  | 46(63) | 27(37) | 8.19 | 2.41 (1.3-4.42) | 0.005<sup>#</sup> |
|  | Agriculture* |  | 46(41.4) | 65(58.6) |  |  |  |
| Occupations<br>of<br>Husband's | Business/ Job |  | 40(48.7) | 38 (51.3) | 0.11 | 1.05(0.53-2.44) | 0.905 |
|  | Labor/Foreign<br>employment |  | 37(48.7) | 39(51.3) |  | 0.95(0.41-2.21) | 0.903 |

|  |  |  |  |  |  |  |
| --- | --- | --- | --- | --- | --- | --- |
|  | Agriculture* | 15(50) | 15(50) |  |  |  |
| Age at marriage | ≥ 20 years | 42(67.7) | 20(32.2) | 11.77 | 3.024(1.59-5.75) | 0.001# |
|  | < 20 Years* | 50(41) | 72(59) |  |  |  |
| Socioeconomic status | Highest quintile | 20(52.6) | 18(47.4) | 0.54 | 1.41 (0.51-3.9) | 0.503 |
|  | Fourth quintile | 19(51.4) | 18(48.6) |  | 1.34 (0.85-3.72) | 0.570 |
|  | Middle quintile | 18(48.6) | 19(51.4) |  | 1.21 (0.43-3.34) | 0.719 |
|  | Second quintile | 24(51.1) | 23(48.9) |  | 1.33 (0.51-3.52) | 0.568 |
|  | Lowest quintile* | 11(44) | 14(56) |  |  |  |
| BMI of respondents | Below normal | 5(45.50) | 6(54.5) | 8.44 | 1.18 (0.33- 4.12) | 0.793 |
|  | Overweight and obesity | 44(63.8) | 25(36.2) |  | 2.49 (1.33- 4.67) | 0.004# |
|  | Normal* | 43(41.3) | 61(58.7) |  |  |  |

\*Reference categories; # significantly associated

The above table showed that, the occupation of respondent was significantly associated with infertility problems among women (p = 0.005). Women engaged in paid job (Business/Job/Labor) were 2.41 times more likely to have infertility problems than women didn’t hve paid job (Housewife/Agriculture) (OR-2.241,95% CI: 1.30-4.42). Age at marriage of women, 20years and above weresignificantly associated with infertility problems in women (p= 0.001).Women married after 20 or above years were 3.02 times more likely to have infertility problem than in women who were married before age of twenty years (OR-3.02, 95%CI: 1.589-5.75).

The socioeconomic status of respondents was not significantly associated with infertility problems. The body mass index of women was significantly associated with infertility problems in women. Leanness/thinness (BMI below18) has no association with infertility where as women with overweight and obesity (>25 BMI) have 2.49 times more likely to have infertility problems than women having normal (18-25) BMI (OR-249, 95% CI: 1.33-4.67).

**Table 6:** Association of infertility problems with other reproductive health problems.

| Independent \ Dependent |  | Dependent<br>n (%) | Non-infertile<br>n(%) | Chi-<br>square<br>value | OR<br>(95%CI) | P value |
| --- | --- | --- | --- | --- | --- | --- |
| Irregular menstruation | Yes | 25(83.3) | 5(16.7) | 15.93 | 6.49(2.35-17.85) | <0.001# |
|  | No* | 67(43.5) | 87(56.5) |  |  |  |
| Ever faced miscarriage or abortion | Yes | 34(60.7) | 22(39.3) | 24.04 | 6.01(2.85-12.67) | <0.001# |
|  | No* | 18(20.5) | 70(79.5) |  |  |  |
| Induced abortion | Yes | 6(28.6) | 15(71.4) | 14.55 | 0.10(0.03-0.35) | <0.001# |
|  | No* | 28 (80.0) | 7 (20.0) |  |  |  |
| VDS | Yes | 14(63.6) | 8 (36.4) | 1.86 | 1.89(0.752-4.74) | 0.173 |
|  | No* | 78(48.1) | 84(51.9) |  |  |  |
| Use of family planning devices before first baby | Yes | 22(84.6) | 4(15.4) | 14.51 | 6.914(2.28-20.99) | 0.001# |
|  | No* | 70(44.3) | 88(55.7) |  |  |  |
| Use of hormonal contraceptives before 1<sup>st</sup> birth | Yes | 13(86.7) | 2(13.3) | 0.12 | 1.44(0.17-12.23) | 0.736 |
|  | No* | 9(81.8) | 2(18.2) |  |  |  |
| Use of family planning devices after first baby | Yes | 24(27.6) | 63(72.4) | 25.15 | 0.025(0.003-0.2) | 0.001# |
|  | No* | 15(93.8) | 1(6.3) |  |  |  |
| Use of hormonal contraceptives after 1<sup>st</sup> birth | Yes | 21(29.6) | 50(70.4) | 0.38 | 1.47(0.43-4.99) | 0.537 |
|  | No* | 4(22.2) | 14(77.8) |  |  |  |

\*Reference catagories; # significantly associated

Binary logistic regression was employed to calculate un-adjusted odds ratio between infertility problems in women and socio-demographic variables. Above table shows that irregular menstruation, history of miscarriage or abortion, induced abortion, use of family planning devices before and after 1^st^ birth of baby was statistically associated with infertility problems in women. Women with irregular menstruation were nearly seven times more likely to have infertility problems than regularly menstruating women (OR-6.49, 95% CI: 2.35-17.85) whereas vaginal discharge syndrome was not significantly associated with infertility problems.

Women who had faced miscarriage or abortion in their reproductive lives found more vulnerable to infertility problems. From the women ever faced miscarriage or induced abortion were six times more likely to have infertility problems than women never faced miscarriage or induced abortion (OR-6.01, 95%CI:2.85-12.67). Although miscarriage or abortion showed association with infertility problems, the induced abortion was protective to infertility problems than women not conducted induction abortion (OR-0.10, 95%CI: 0.03-0.35). Similarly use of family planning devices before first baby was significantly associated with infertility problems in women. The women who used any family planning devices before birth of first baby were nearly seven times more likely to have infertility problems than women who didn’t used any family planning devices before first baby (OR-6.914, 95%CI: 2.28-20.99). The use of major types of contraceptives (hormonal/non-hormonal) by women before first baby by women was not significantly associated with infertility problems but women who used hormonal family planning devices before birth of first baby found 1.444 times more likely to have infertility problems than women who had used non-hormonal family planning devices before first baby (OR-1.44, 95%CI: 0.17-12.23). Use of family planning devices after birth of first baby was statistically associated with infertility problems in women (OR-0.025, CI: 0.003-0.20). Use of hormonal contraceptives after birth of baby was 1.470 times more likely to have infertility problems in women than women who used non-hormonal contraceptives but the difference was not significant (OR-1.47, 95% CI: 0.43-4.992).

#### 4.3.2 Association of infertility with frequency of visit for treatment and cost for treatment of RH problems

**Table 7:**
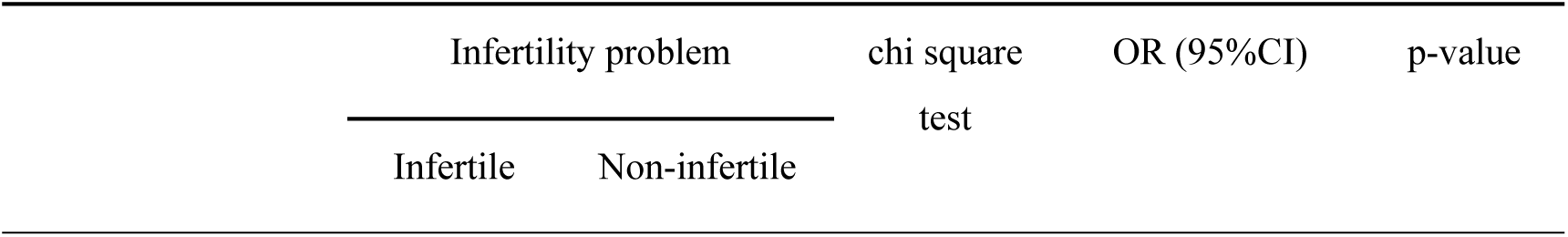
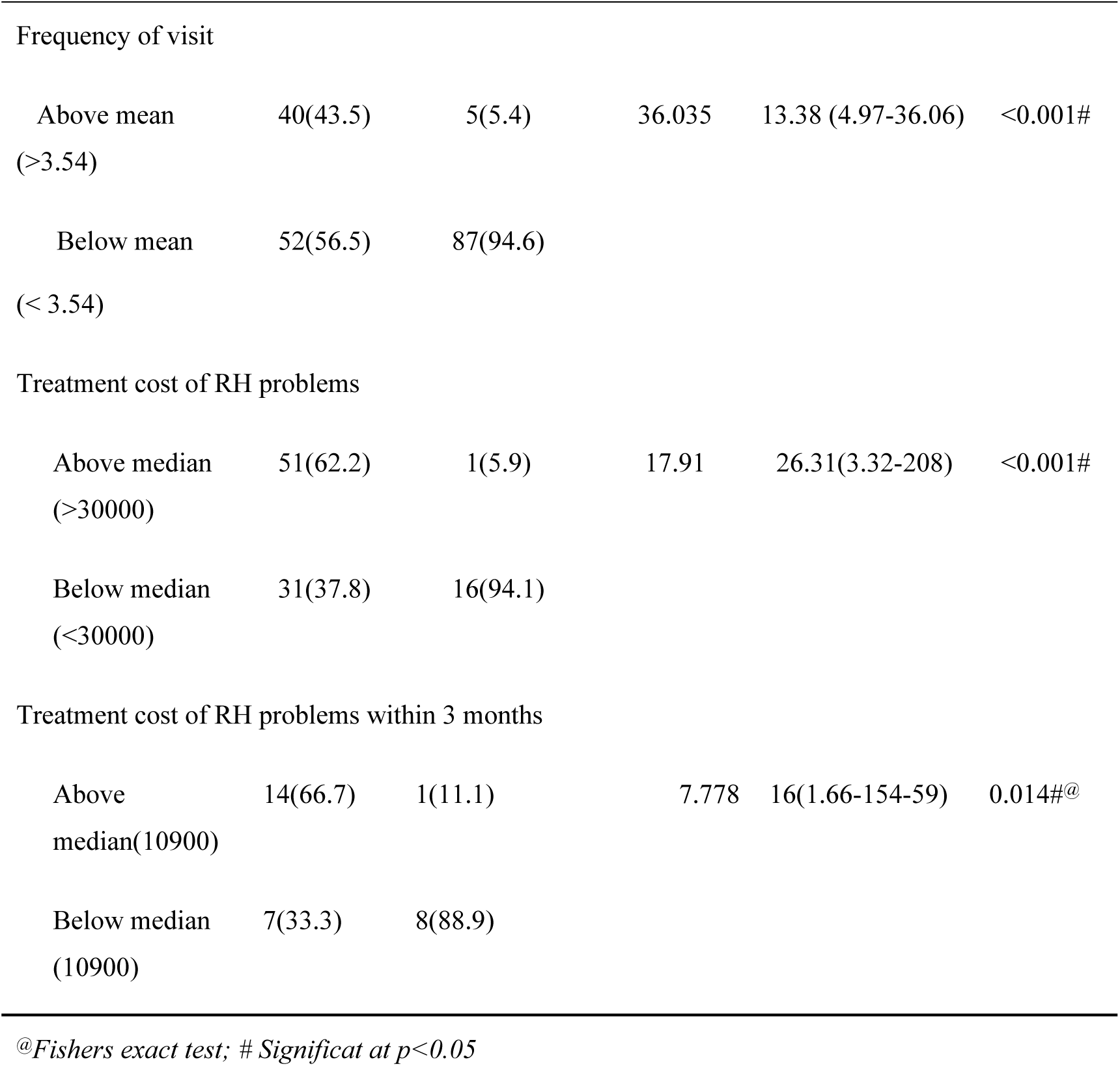
Association of infertility with frequency of visit for treatment and cost for treatment of RH problems.

| Infertility problem |  | chi square<br>test | OR (95%CI) | p-value |
| --- | --- | --- | --- | --- |
| Infertile | Non-infertile |  |  |  |

|  |  |  |  |  |  |
| --- | --- | --- | --- | --- | --- |
| Frequency of visit |  |  |  |  |  |
| Above mean<br>(>3.54) | 40(43.5) | 5(5.4) | 36.035 | 13.38 (4.97-36.06) | <0.001# |
| Below mean<br>(< 3.54) | 52(56.5) | 87(94.6) |  |  |  |
| Treatment cost of RH problems |  |  |  |  |  |
| Above median<br>(>30000) | 51(62.2) | 1(5.9) | 17.91 | 26.31(3.32-208) | <0.001# |
| Below median<br>(<30000) | 31(37.8) | 16(94.1) |  |  |  |
| Treatment cost of RH problems within 3 months |  |  |  |  |  |
| Above<br>median(10900) | 14(66.7) | 1(11.1) | 7.778 | 16(1.66-154-59) | 0.014#@ |
| Below median<br>(10900) | 7(33.3) | 8(88.9) |  |  |  |

@Fishers exact test; # Significat at $p < 0.05$

Women with infertility problems were 13.38 times more likely to visit different places for treatment for more than 3 times than non-infertile women with reproductive problems (OR-13.38, 95%CI: 4.97-36.06). The infertile women found investing significantly more amount of money than non-infertile women (OR-26.31, 95%CI: 3.32-208). Similarly the amount of cost invested for treatment of RH problems was significantly different between infertile & non-infertile women (OR-16, 95%CI: 1.66-154-59)

#### 4.3.3 Association of psychosocial variables with infertility problems

**Table 8:**
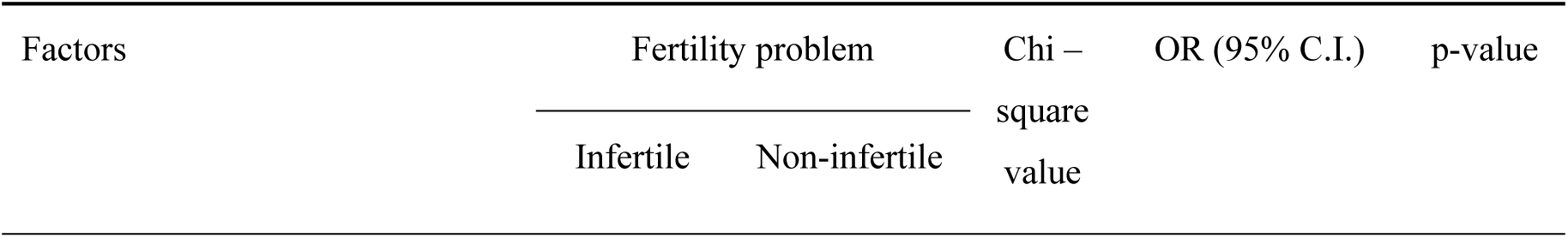
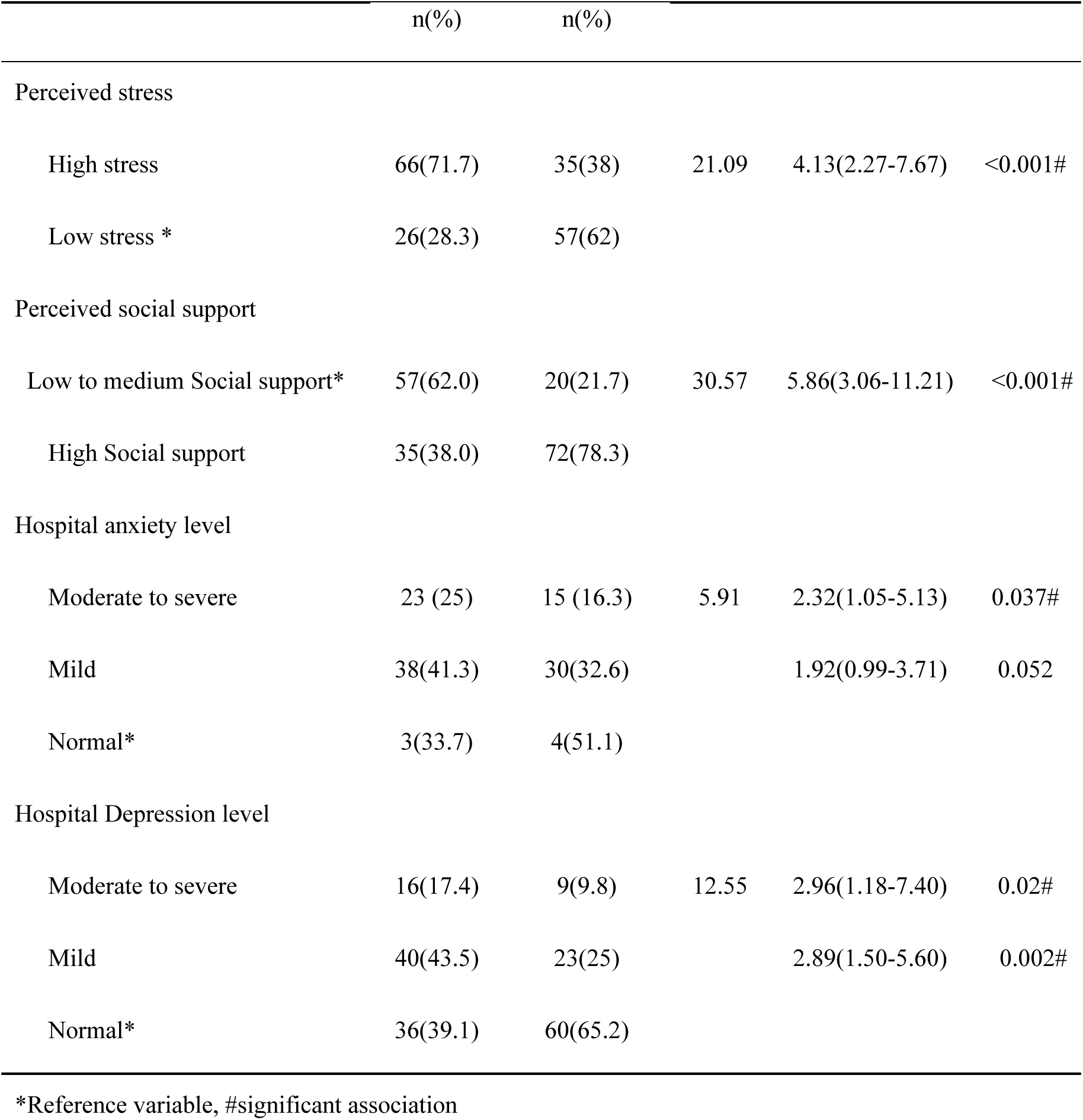
Association of perceived stress, anxiety and depression levels, and perceived social support with infertility problems.

| Factors | Fertility problem |  | Chi – square value | OR (95% C.I.) | p-value |
| --- | --- | --- | --- | --- | --- |
|  | Infertile | Non-infertile |  |  |  |

|  | n(%) | n(%) |  |  |  |
| --- | --- | --- | --- | --- | --- |
| Perceived stress |  |  |  |  |  |
| High stress | 66(71.7) | 35(38) | 21.09 | 4.13(2.27-7.67) | <0.001# |
| Low stress * | 26(28.3) | 57(62) |  |  |  |
| Perceived social support |  |  |  |  |  |
| Low to medium Social support* | 57(62.0) | 20(21.7) | 30.57 | 5.86(3.06-11.21) | <0.001# |
| High Social support | 35(38.0) | 72(78.3) |  |  |  |
| Hospital anxiety level |  |  |  |  |  |
| Moderate to severe | 23 (25) | 15 (16.3) | 5.91 | 2.32(1.05-5.13) | 0.037# |
| Mild | 38(41.3) | 30(32.6) |  | 1.92(0.99-3.71) | 0.052 |
| Normal* | 3(33.7) | 4(51.1) |  |  |  |
| Hospital Depression level |  |  |  |  |  |
| Moderate to severe | 16(17.4) | 9(9.8) | 12.55 | 2.96(1.18-7.40) | 0.02# |
| Mild | 40(43.5) | 23(25) |  | 2.89(1.50-5.60) | 0.002# |
| Normal* | 36(39.1) | 60(65.2) |  |  |  |

\*Reference variable, #significant association

The infertility problem was significantly associated with different psychosocial factors. The infertility problem was significantly associated with perceived stress (pvalue < 0.001). Women with infertility problems have more than four times more likely to perceived high stress then non-infertile women (OR-4.134, 95%CI: 2.27-7.67). Similarly women with infertility problems have 5.86 times more likely to perceive low social support than non-infertile women (OR-5.863, 95%CI: 3.06-11.21).

The infertile women were 2.32 times more likely to have moderate to severe level of anxiety than non-infertile women. Similarly the mild level of anxiety was higher among infertile women then non-infertile (OR-1.920, 95%CI: 0.993-3.713). Infertile women were nearly three times more risk to moderate to severe level of depression (OR-2.96, 95%CI: 1.18-7.40) and mild level of depression (OR-2.89, 95%CI: 1.50-5.60) than non-infertile women.

**Table 9:**
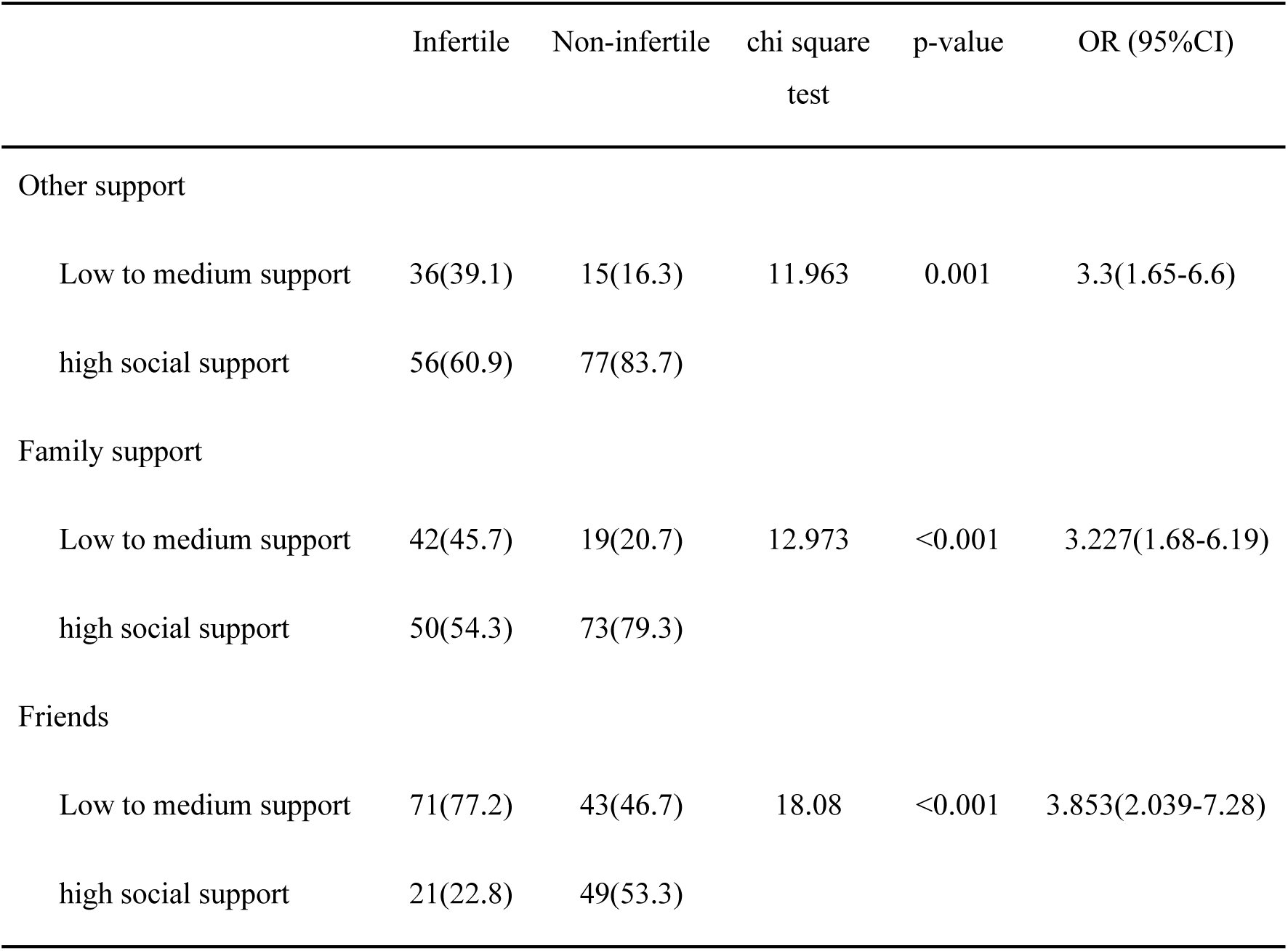
Association between infertility problems with perceived social support.

|  | Infertile | Non-infertile | chi square<br>test | p-value | OR (95%CI) |
| --- | --- | --- | --- | --- | --- |
| Other support |  |  |  |  |  |
| Low to medium support | 36(39.1) | 15(16.3) | 11.963 | 0.001 | 3.3(1.65-6.6) |
| high social support | 56(60.9) | 77(83.7) |  |  |  |
| Family support |  |  |  |  |  |
| Low to medium support | 42(45.7) | 19(20.7) | 12.973 | <0.001 | 3.227(1.68-6.19) |
| high social support | 50(54.3) | 73(79.3) |  |  |  |
| Friends |  |  |  |  |  |
| Low to medium support | 71(77.2) | 43(46.7) | 18.08 | <0.001 | 3.853(2.039-7.28) |
| high social support | 21(22.8) | 49(53.3) |  |  |  |

The above table showesthat, family, friends and other subscales of perceived social support were statistically associated with infertility problem (p<0.001). The women with infertility problems perceived lower friend, family and other social support than non-infertile women.

#### 4.3.4 Factors associated with Quality of life

**Table 10:** Quality of life score mean difference among case and control.

| Quality of life | Infertile |  |  | Non-infertile |  |  | Mean Difference | p value |
| --- | --- | --- | --- | --- | --- | --- | --- | --- |
|  | N | Mean | SD | N | Mean | SD |  |  |
| Physical Health | 92 | 60.66 | 9.38 | 92 | 68.64 | 11.16 | -7.98 | < 0.001 |
| Psychological | 92 | 50.58 | 9.92 | 92 | 57.64 | 9.61 | -6.79 | < 0.001 |
| Social Relations | 92 | 57.9 | 12.04 | 92 | 67.75 | 13.37 | -9.78 | < 0.001 |
| Environment | 92 | 48.6 | 10.37 | 92 | 57.98 | 10.4 | -9.34 | < 0.001 |
| Overall QOL | 92 | 54.0 | 7.79 | 92 | 62.3 | 7.82 | -78.28 | < 0.001 |

In this case control study, 184 infertile and non-fertile women with the mean age 32.86±6.201 were compared. The mean score of overall quality of life and it all physical, psychological, social relationship and environmental domains in infertile women was significantly lower than that of non-infertile women (p= < 0.001).

**Table 11:**
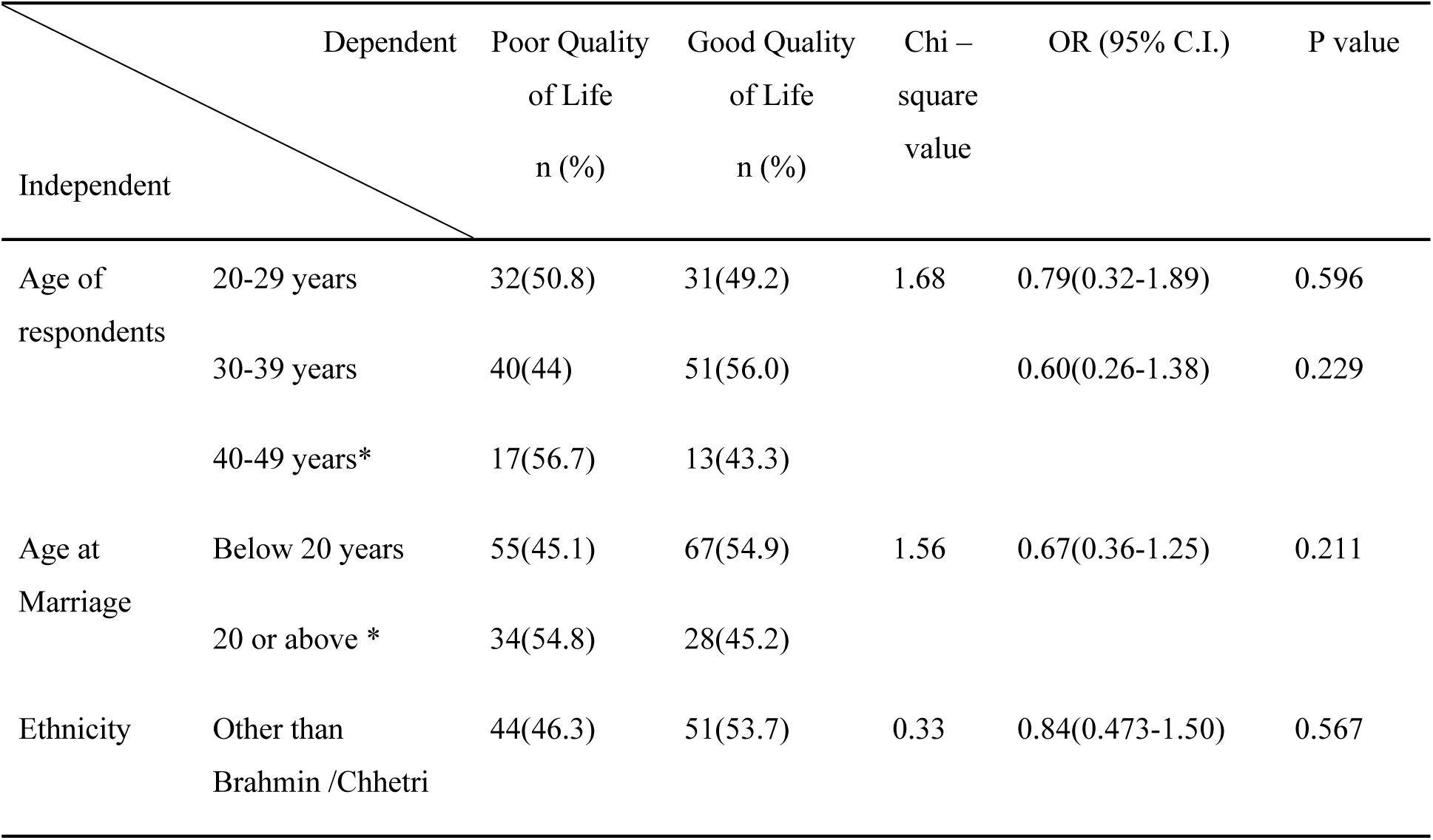
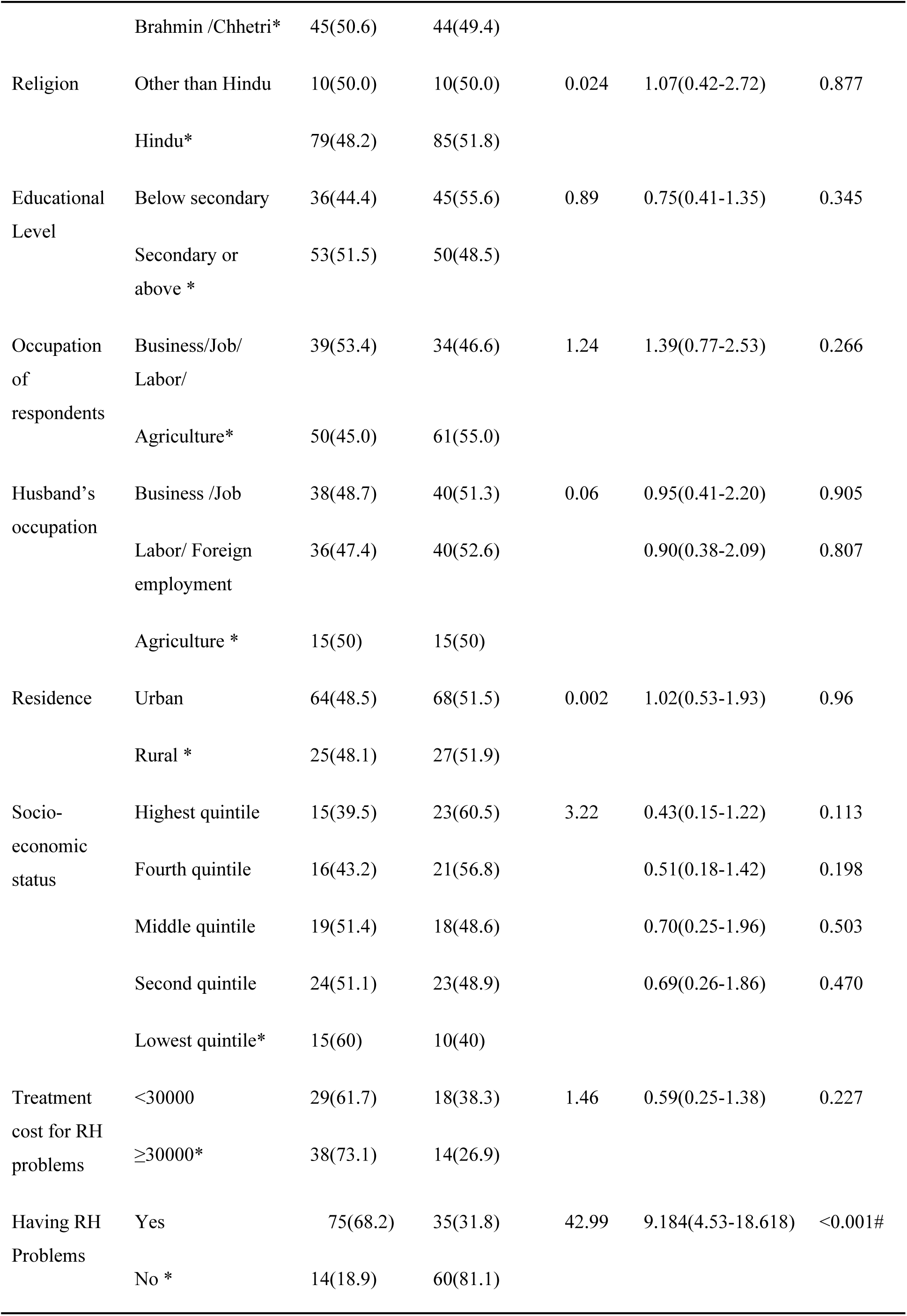
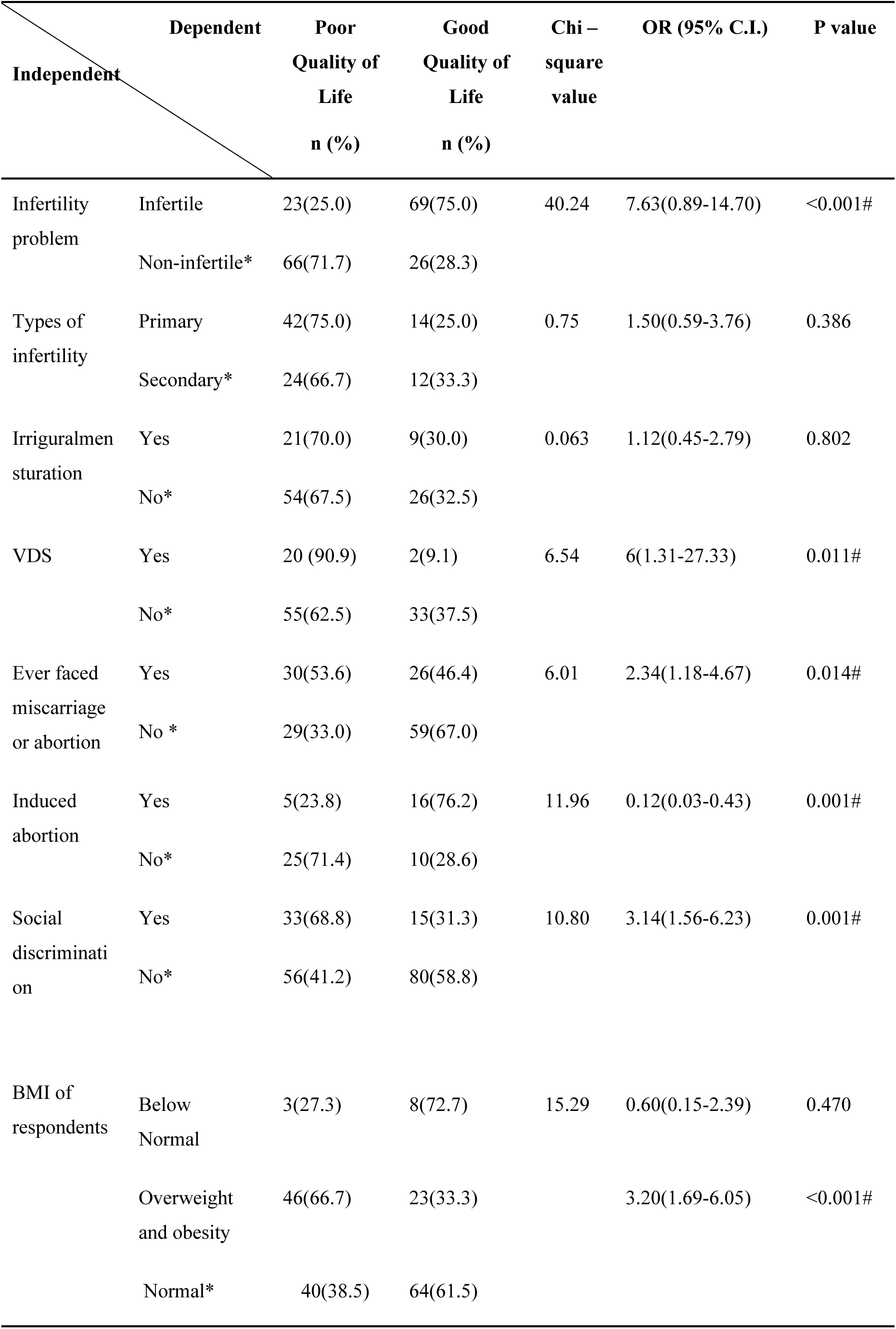
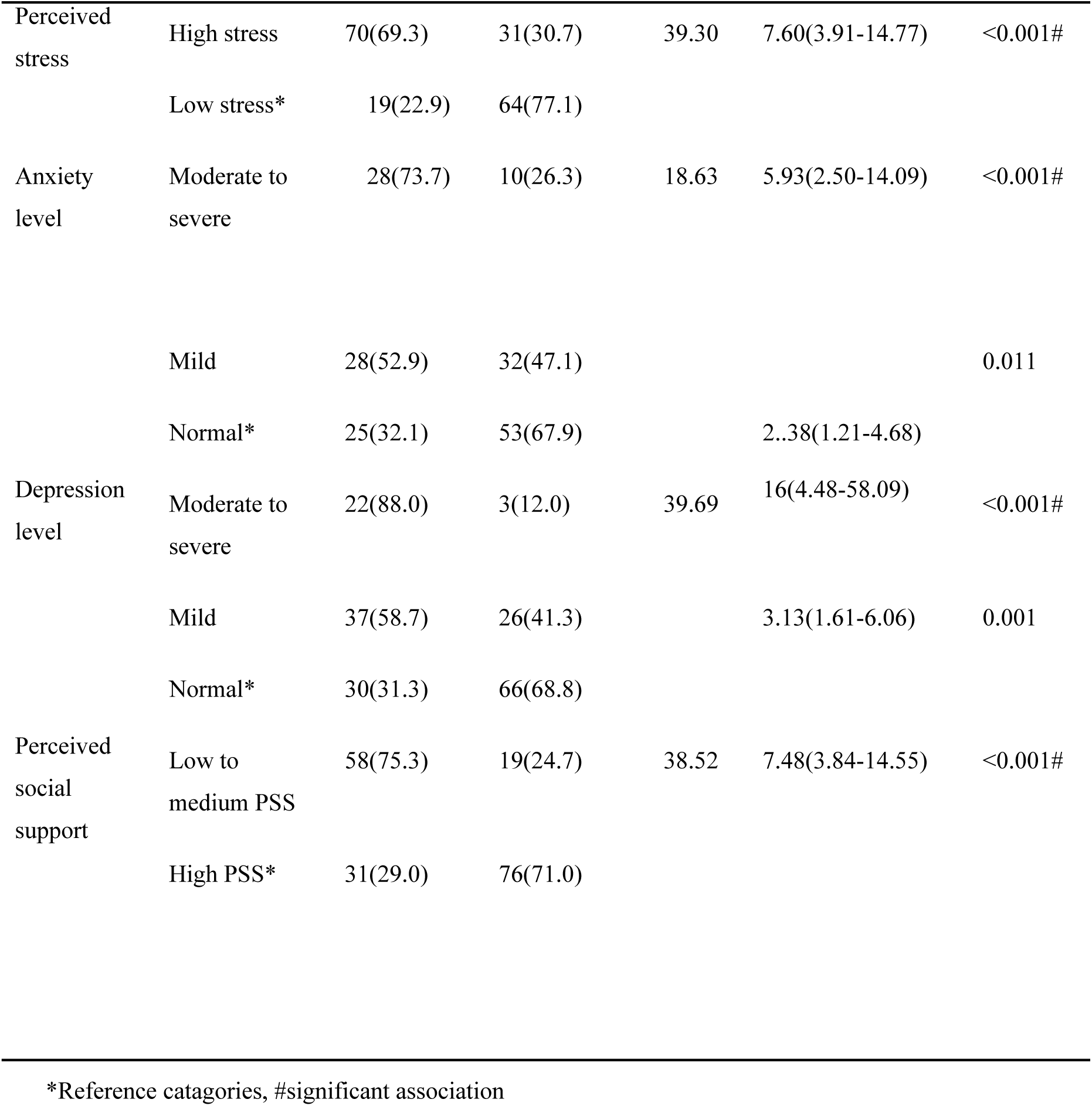
Factors associated with overall quality of life.

The quality of life score by WHO-QoL-BREF was categorized into two groups based on mean score (52.01±7.98). Respondents achieved average quality life score less than mean score were categorized as poor quality of life and greater or equal mean score as good quality of life.

Binary logistic regression was used to calculate the crude odds ratio between overall quality of life and socio-demographic variables. Higher the socioeconomic status the quality life found increased, but the association was not statistically significant. Women in highest quintile as per IWI group were more protective to have poor quality of life than women in lowest quintile of IWIgroup (OR-0.435, 95%CI: 1.55-1.219). Similarly women in fourth quintile group were also found protective to have poor quality of life than women in lowest quintile of IWI group (OR-0.508, 95%CI: 0.181-1.424).

Among different reproductive health related factors, having RH problems, infertility problems, having VDS, and social discrimination were significant for poor quality of life. Having RH problems (OR-9.184, 95%CI: 4.53-18.618), infertility problems (OR-7.63, 95%CI: 3.89-14.70), VDS (OR-6, 95%CI: 1.317-27.33), ever faces miscarriage or abortion (OR-2.34, 95%CI: 1.18-4.67) social discrimination (OR-3.14, 95%CI: 1.56-6.23) were more likely to have poor quality of life than those who have no such reproductive problems. Where as having history of induced abortion found protective for poor quality of life then women without history of induced abortion (OR-0.12, 95%CI: 0.036-0.433).

Similarly the poor quality of life was significantly associated with different anthropometric and psychosocial variables such as BMI of respondents, perceived stress, anxiety levels, depression levels and perceived social support by respondents. The women with overweight and obesity were three times more likely to have poor quality of life than women with normal BMI (OR-3.20, 95%CI: 1.691-6.054). Similarly women with high perceived stress were 7.60 times more likely to have poor quality of life than women with low perceived stress (OR-7.60, 95%CI: 3.91-14.77). The level of anxiety and depressions were significant for poor quality of life. Moderate to severe level of anxiety (OR-5.936, 95%CI: 2.500-14.092), mild anxiety level (OR-2.38, CI: 1.21-4.67), moderate to severe level of depression (OR-16, 95%CI: 4.48-58.09), mild depression level (OR-3.13, 95%CI: 1.61-6.06) levels of depression and anxiety were more likely to have poor quality of life than those with normal anxiety and depression levels. Likewise the women who perceived low social supports were 7.484 times more likely to have poor quality of life than perceived high social supports (OR-7.48, 95%CI: 3.84-14.56).

#### 4.3.5 Factors associated with quality of among infertile women

**Table 12:**
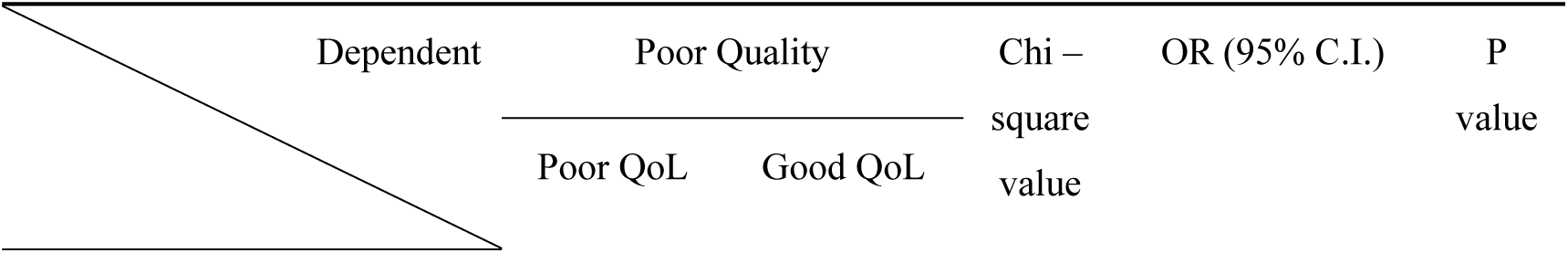
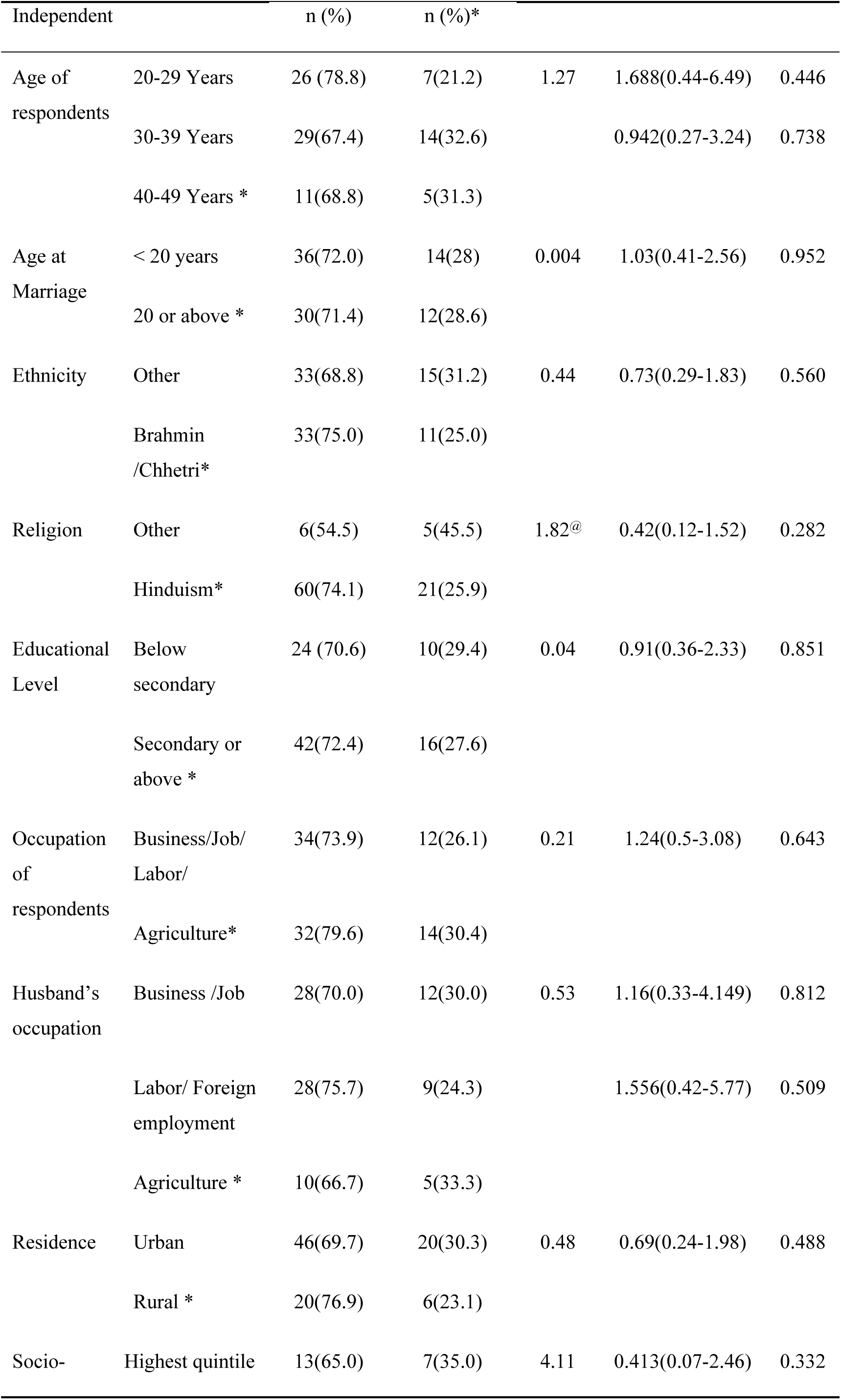
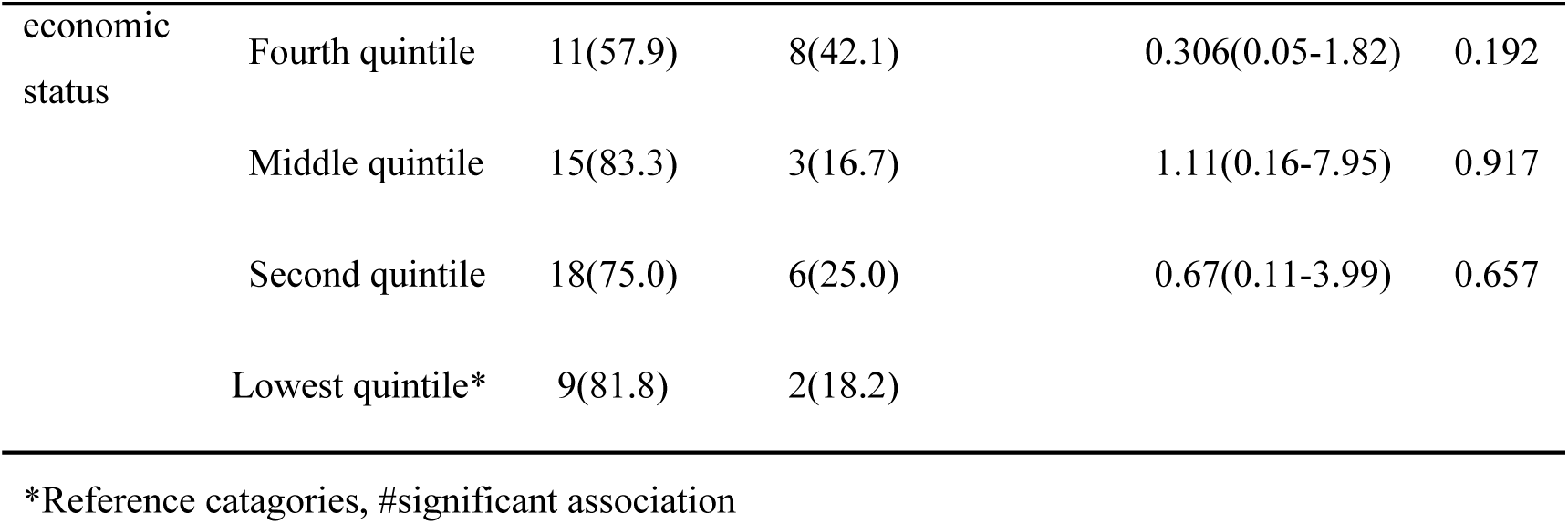
Associated between socio-demographic and quality of life of infertile women.

Above table shows that none of the socio-demographic characteristics were significantly associated quality of life of infertile women.

## DISCUSSION

### 5.1 Summary of the Discussion

The findings of the study show that the quality of life among women with infertility was poor than that of non-infertile women. High perceived stress, mild to severe level of depression, perceived social support and having reproductive health problems were found significant determinates of QoL among married women of reproductive age. Among infertile women, perceive stress and social support were significant determinants of QoL whereas level of depression, social support and RH problems were found significant determinates of QoL among non-infertile women.

### 5.2 Socio-Demographic Characteristics

The main objective of this research was to assess the quality of life among infertile and non-infertile women and to identify factors associated with quality of life to them. Infertility is highly stressful to married couple and influences several aspects of life in women. It adversely affect the mental and social health of infertile couples.^2^ It is considered reproductive as well as social problem. ^3,4^Although the institutional based study on infertility and its impact on quality of life have been widely studied, there are relatively few community based studies of infertility and its impact on quality of life.

In this study 123 couples with infertility problem were screened through community based survey among 1351 married women of reproductive age (20-49). The prevalence of infertility problems found 9.1%, it was reported 5.4% by a study conducted in Eastern Nepal.^5^ Similarly a gynecological camp conducted in Bajhang district has also showed 14.2% women had sub-fertility problems.^6^ Survey conducted in eight districts of Nepal by UNFPA and IOM in 2006 found 7.4% prevalence of infertility among reproductive age women.^7^ From this study and other different studies showed nearly one in ten couple of reproductive aged have infertility/subfertility problems problem in Nepal.

In this study, 184 infertile and non-infertile women with the mean age 32.86±6.2 and mean age at marriage 18.70±2.79 years were compared. The mean age at marriage (19.35) among infertile women was more than that of non-infertile women, but the different was not significant (p=0.21). Similarly case control study conducted in rural Northern China has shown that the mean age at marriage among infertile women was significantly higher than that of non-infertile women.^8^

The median time of willingness to give birth to new baby was 36 months (minimum-o, maximum-300, IQR-96). Similarly mean duration of infertility was (2.92±2.25) years^9^ and (4.3±0.5) years,^10^ but somewhat longer duration (7.4±5.2 years) has shown by another study.^11^

The causes of infertility were due to of male factors (16.36%) and female factors (29%). In 11.82% both male and female factors were observed and 42.73% of couple had unexplained infertility. More than half of infertility was secondary infertility 56.5%. Similarly the proportion of secondary infertility cases shown by study conducted in Eastern Nepal was also 56.5%.^5^whereas a cross-sectional study in china showed nearly similar prevalence of infertility (13.09%) like our study but the proportion of primary(7.58%) and secondary(92.42%) infertility was different.^8^

A hospital based study at Dhulikhel Nepal in 2019 has also showed that nearly half 48.8% infertility cases were female factor infertility, 23.9% male factor, 26.6% both factor and 14.4% unexplained factor infertility. The study in Dhulikhel hospitalsh has shown that, primary infertility were three times more common than secondary infertility,^11^ but in our community based study the secondary infertility found more than half (56.5%) of the cases. We found that faith healer was first contact point for treatment of infertility of 32.9% infertile women, home/using herbal medicine (29.3%), Private hospitals (26.1%) and others. In this study the average frequency of visiting different place for treatment of infertility problems by women with primary infertility (6.11 times) was nearly two times more than that of secondary infertility (3.33) and difference was significant (p=0.027). Which could due to of more family and social pressure to primary infertile women than secondary infertility or taking infertility more seriously by primary infertile women than women with secondary infertility.

### 5.3 Factors associated with infertility

There are verities of factors affecting fertility. In this study the demographic and reproductive health related factors were studied. The different factors, occupation, age at marriage, and BMI were significantly associated with infertility. The women engaged in business/job found 2.41times more risk of infertility than women engaged in agriculture (OR-2.241, 95%CI: 1.3-4.42). The delay marriage (after 20 years) found also associated with infertility (p=<0.001).

With increasing sedentary lifestyle, number of overweight and obesity is also increasing, which is important factors leading infertility. In this study the body mass index of women was significantly associated with infertility, both women with underweight and overweight found at risk of infertility than normal weight women. Whereas the risk of infertility in women with underweight was not significant (OR-1.18, p=0.79) but women with overweight and obesity (> 25 BMI) found 2.49 times at risk if infertility than normal weight women (BMI 18.5-25) (OR-2.49, p=0.004). Similarly this study is consistent with the study conducted in china that revealed underweight and obese women had high incidences of infertility, and the incidence of infertility was highest in the obesity group.^8^

Among different reproductive health related factors, irregular menstruation, history of miscarriage or abortion, induced abortion, and use of family planning devices before and after 1^st^ birth found significantly associated with infertility. Women with irregular menstruation found 6.49 times more risk of infertility than regularly menstruating women (OR-6.49, 95%CI: 2.235-17.85). A case control study has also showed that 2.29 times more risk of infertility among women with irregular menstruation (OR-2.29), similarly women having had pelvic procedure or induced abortion(MVA) was significantly associated with infertility.^12^

In multivariate logistic regression analysis, five factors were independent and three determinants, significantly associated with infertility were: occupation (AOR-16.88, 95%CI: 1.149-191.58), use of family planning devices before 1^st^ child (AOR-16.59, 95%CI: 1.42-194.5) and induced abortion (AOR-0.086, 95%CI: 0.01-0.969). In our study induced abortion showed protective role for infertility.

### 5.4 Perceived Stress

In the current study we found that only occupation of respondent, BMI, infertility problems and social discrimination & violence were significantly associated with perceived stress. Women engaged in paid job found perceived more stress than women engaged in agriculture (OR-2.11, p=0.017). Similarly women with overweight and obesity also found perceived more stress than normal weight women (OR-2, p=0.03). This may be due to of body image dissatisfaction of obese women.

This study revealed that, infertile women were four times more risk of perceived high stress (OR-4.13, p=<0.001) and perceived social discrimination (OR-4.03, p=<0.001) than non-infertile women. In line with this study, People facing infertility problems have higher risk of anxiety, stress, and depression.^4,13–16^ Similarly a clinic-based study in India revealed that, 80% Prevalence of stress among infertile women,^17^ a Similar study has shown 19% moderate and 13% prevalence of severe depression among infertile women.^14^

A study was conducted in India Age, Marital years, duration of infertility, male factor infertility, history of gynecological surgery, ovulation induction, presenting psychiatric morbidity, infertility related coping difficulty were signifactly associated with infertility related stress.^17^ Study by Wiwekok et al. 2017 showed different level of stress due to of infertility and stress was significantly associated with duration of infertility experienced by patient (p < 0.05).^18^

A cross sectional study among infertile women visiting, Tokai University Hospital found that anxiety and depression among infertile women were associated with felling stress and support by their husbands (Matsubayashi et al., 2004). Different studies in the field of infertility and reproductive health problems have verified that the different factors like age at diagnosis, cause of infertility, duration of infertility, coping abilities, treatment cost, treatment failures, psychiatric morbidity, psychosocial support, stigma, and discrimination are mostly related to infertility-specific stress among infertile people. ^4,17,19,20^

### 5.5 Anxiety

Mean score of the anxiety of infertile women was 8.71± 3.05where a study was conducted in Nederland showed that the mean score of anxiety level was (5.6±3.9SD) among in fertile women.^21^

This study was found that 57.61 percent of the women reported low to severe level depression among 184 women. In one of the largest studies to date, 352 women were assessed in infertility clinics in northern California. It was determined that 76% of the women reported significant symptoms of depression.^22^ In another recent study of 174 women undergoing infertility treatment, 39% met the criteria for major depressive disorder.^23^

This study was found that the low relationship between age and anxiety level. But another study conducted in Hungary at 2014, doesn’t support to this which was significant at (p-value=0.026).^24^

This study revealed that, socioeconomic status and infertility were significantly associated with moderate to severe level of anxiety. Increasing socioeconomic status found protective to moderate to severe level of anxiety. This may be due to of socially and economically secure feeling of high socioeconomic status women. The infertile women were 2.233 times more risk of moderate to severe level of anxiety than non-infertile women(OR-2.323, p=0.037). Some studies have indicated that social support is related to lower depression and anxiety level.^24^

### 5.6 Depression

This study was found that man score of the depression of infertile women was 8.14±2.67, where as similar type of study was conducted in Nederland showed that the mean score of depression level was 3.6 (3.3SD) among in fertile women.^21^

This study was found that 47.82 percent of the women reported low to severe level depression among 184 women. Study of 352 women in infertility clinics in northern California determined that 56% of the women reported significant symptoms of depression.^22^

Multinomial logistic regression was employed to analyzed factors associated depression, which found socioeconomic status and infertility problems were significantly associated with level of depression. The age and educational status of women werenot significant with level of depression, but similar type of studies showed that age (p-value=0.018).^25, 26^

This study revealed that, socioeconomic status and infertility and types of infertility were significantly associated with moderate to severe level of depression. Increasing socioeconomic status found protective to moderate to severe level of depression. This may be due to of socially and economically secure feeling of high socioeconomic status women. The infertile women were 2.96 times more risk of moderate to severe level of depression than non-infertile women(OR-2.96, p=0.02). Similarly women with primary infertility found 21 times more risk of severe to moderate level of depression than women with secondary infertility. Which is supported by frequency of visit health facility by secondary infertility is lower than primary infertility. This could be due to of feeling more satisfied with presence of child among secondary infertile women than primary infertile women.

Infertility can cause psychological distress and has a negative impact on quality of life. A cross sectional study conducted Iran also showed that both males and females’ depression exuded an actor effect on their own QoL (β = − 0.589, p < 0.001; β = − 0.588, p < 0.001, respectively).^25^

### 5.7 Perceived Social support

Social support is a source of coping it has great importance for the infertile woman to help preserve her physical and mental health. This study showed that the mean score of total family support among infertile women (58.77±7.87) was significantly lower than that of non-infertile women (65.67±7.78) (p<0.001). Similarly the subscale wise average score of social support were also significantly lower in infertile women than non-infertile women. A case control study conducted among infertile and fertile women showed that the total mean score as well as subscale wise mean score of perceived social support among infertile women was significantly higher than fertile women(p=0.001).^3^

This study revealed that women with infertility problems have 5.862 times more likely to perceive low social support than non-infertile women (OR-5.863, 95% CI: 3.060-11.211). Although the association between types of infertility and perceived social support was not statistically significant, the women with primary infertility problems have 1.89 times more likely to have low social support than women with secondary infertility problems (OR-1.89, 95%CI: 0.80-4.47). The respondents who perceived social discrimination and violence were 3.56 times more likely to have less social support than not perceived social discrimination (OR-3.568, 95%CI: 1.790-7.113). Similarly women with below secondary level educational status have little more likely to perceive high social support than women studied secondary and above (OR-1.188, 95%CI: 0.65-2.14). The women with high stress found 4times more likely to have low perceived social support than women with low stress (OR-4.08, 95%CI: 2.155-7.728). A cross section study also revealed that infertility stress was strongly associated with perceived social support in both man and women partner of infertile couple. MSPSS statistically negative significant relationship was found between the scales at the level of p<0.01.^26^

### 5.8 Quality of life

.The aim of this study was to determine and compared the QoL and its related factor among infertile and non-infertile women using WHOQoL-BREEF questionnaire. In this case control study the mean score of quality of life of total infertile and non - infertile women was 58.15±8.28. The domain wise and overall mean score of quality of life found significantly lower in infertile women(54±7.79) than non-infertile women(62.3±7.82) (P=<0.001). In line with our findings mousavai et al. 3013, revealed that infertility had negative effect on quality of life of couple.^9^Similarly another studies have also showed infertile women experienced lower quality of life.^24,27^

Similarly a previous case control study conducted in hospital seating in Nigeria had shown that lower mean score of overall Qol among infertile than fertile women but the difference was not significant (0.095), the mean score of physical and psychological domain were higher among infertile women.^28^

Another case control study among 50 infertile and 50 fertile women has shown dissimilar findings than our study where the mean score of physical, psychological, relationship and QOL in infertile women was significantly more than fertile women, although the mean score of environmental health in infertile women was more than fertile ones, but the difference was not significant (p= 0.15).^3^ Other studies have shown that infertility is a distressing and painful experience, especially for women and is associated with lower quality of life scores. ^8,29,30^

### 5.9 Factors Associated with Quality of Life

One of the objectives of this study was to identify the factors associated with quality of life of infertile women. In this study the average quality of life of infertile women found poorer than non-infertile women. The different demographical, psychosocial, and reproductive health related factors were analyzed in order to identify the potential determinants factors of quality of life. Among different factors the presence of RH problems, infertility, VDS, ever faced miscarriage or abortion, social discrimination, overweight and obesity, perceived stress, level of anxiety, level of depression, and perceived social support were significant with overall quality of life of women during bivariate analysis. From multivariate analysis only having RH problems (AOR-4.06, p=0.001), moderate severe level of depression (AOR-21.11, p=0.005), mild level of depression (AOR-2.41, p=0.0055), perceived social support (AOR-3.14, p=0.005) and perceived stress (AOR-2.34, p=0.004) found significant determinants of quality of life of women.

The quality of life of infertile women found associated with different psychosocial and reproductive health related problems. Overweight and obesity (BMI>25) in infertile women found associated with poor quality of life of infertile women (p=0.016). Infertile women with overweight were more than three times risk of poor quality of life than normal weight (BMI-18-25) infertile women (OR-3.456, 95%CI: 1.25-9.52).

In this study the perceived stress found significantly associated with quality of life of infertile women (p=< 0.001).infertile women with high perceived stress were 8.96 time more likely to have poor quality of life (OR-8.96, 95%CI: 3.17-25.29). Consistent with these results, a previous study had shown that infertile women experience more feelings of helplessness in comparison to fertile women and they are more at risk of mental and emotional disorders, depression, anxiety, low self esteem and marital dissatisfaction.^31^

The results showed that women with a high anxiety and depression had lower levels of QoL. Among infertile women, moderate to severe level of anxiety (OR-6.632, 95%CI: 1.312-33.512), moderate to severe level of depression (OR-9.545, 95%CI: 1.123-80.506) levels of depression and anxiety were more likely to have poor quality of life than those with normal anxiety and depression levels which is in line with the results of other authors.^4,32,33^ A study conducted infertility center in Tehran, has also found that poor quality of life of women facing infertility problems and quality of life is found associated with high depression and anxiety level, failure in previous treatment and unknown cause of infertility. Multivariate analysis showed the anxiety (β = -1.59, p < 0.001) & depression (β = -2.09, p < 0.001) had a negative impact on QoL (MaroufizadehGhaheri and Omani Samani, 2017).^4^

The perceived social support among infertile women found significantly lower than that of non-infertile women (P< 0.0001). Infertile women were less supported than fertile ones by society, family, friends and other people. The infertile women who perceived low social supports were 3.96 times more likely to have poor quality of life than perceived high social supports (OR-3.96, 95%CI: 1.53-10.64). Another study showed that A study conducted infertility center in Tehran, has also found that poor quality of life of women facing infertility problems and quality of life is found associated with high depression and anxiety level, failure in previous treatment and unknown cause of infertility. Multivariate analysis showed that the anxiety (β = -1.59, p < 0.001) and depression (β = -2.09, p < 0.001) had a negative impact on QoL.^4^ Similarly another study revealed that infertility reduces the mental and physical health, social relationship and quality of life.^3^

From this study, we found that QoL was significantly associated with Body mass index level; depression and anxiety level in bivariate analysis, whereas this relationship was not observed in multivariate analysis after adjusting other variables. From multiple logistic regression only two factors, Perceived stress (AOR-10.13, 95%CI: 3.52-29.18) and perceived social support (AOR-3.412, CI: 1.15-10.101) found as important determinants of quality of life among infertile women. In Nepali context more priority is given to offspring’s, and has cultural. Family, and power related value, child are assumed to be power of family and representative of father and family so family and psychosocial pressure may be more in infertile couple, especially women faced social discrimination and violence, which would have had negative impact on their quality of life.

## CHAPTER-VI-SUMMARY, CONCLUSIONS & RECOMMENDATIONS

### 6.2 Conclusions

Prevalence of infertility found one in ten among married women of reproductive age group in Gandaki province. Overall as well as inter-domain quality of life was significantly lower in infertile women as compared to non-infertile women. Majority of the cases found that perceived a social discrimination and violence due to RH problems. BMI, ever faced miscarriage, social discrimination, Perceived stress, anxiety, depression, and social support were found significantly associated with quality of life of infertile women. From multivariate analysis, Perceived stress, and perceived social support found as important determinants of quality of life among infertile women and among non-infertile women moderate to severe level of anxiety, depression, perceived social support and having RH problems found as determinants of quality of life among non-infertile women.

### 6.3 Recommendations

The aim of this study is to empower decision makers to design evidence based appropriate and necessary measures that can promote overall reproductive health of people. So findings and recommendations from this study could be valuable for different concerned bodies which could further be beneficial in improving program implementation modality in near future for country like Nepal by developing a research questions for large scale studies to identify factors associated with infertility ans its psychosocial impacts in individuals, family and community.

## Data Availability

Corresponding author will provide all necessary data upon the request

